# A structural equation modelling approach to understanding the determinants of childhood vaccination in Nigeria, Uganda and Guinea

**DOI:** 10.1101/2022.10.26.22281554

**Authors:** James Bell, Belinda Lartey, Marcos Fernandez, Natasha Darrell, Holly Exton-Smith, Cassie Gardner, Emily Richards, Abolaji Akilo, Emmanuel Odongo, James Ssenkungu, Rigobert Kotchi Kouadio, Mamadi Cissé, Axel Bruno Ayiya Igowa Rérambyah, Maikol Adou, Rebecca West, Sunny Sharma

**Affiliations:** Ipsos Healthcare, London, UK; Ipsos Nigeria, Lagos, Nigeria; Ipsos Uganda, Kampala, Uganda; Ciblage, Dakar, Senegal; Ciblage, Conakry, Guinea; Ciblage, Abidjan, Ivory Coast; Boston University School of Public Health, Boston, United States

## Abstract

Vaccines have contributed to reductions in morbidity and mortality from preventable diseases globally, but low demand for vaccination threatens to reverse these gains. Explorations of the determinants of vaccination uptake may rely on proxy variables to describe complex phenomena and construct models without reference to underlying theories of vaccine demand. This study aimed to use the results of a formative qualitative study (described elsewhere) to construct and test a model to explain the determinants of vaccination uptake. Using the results of a survey among more than 3,000 primary caregivers of young children in Nigeria, Uganda and Guinea, factor analysis produced six explanatory factors. We then estimated the effects of each of these factors on uptake of immunization using a structural equation model. The results showed that the probability that a child is fully vaccinated increases if a caregiver has support from others to vaccinate them (B= 0.33, β= 0.21, p<0.001) and if caregivers had poor experiences with the healthcare system (B= 0.09, β= 0.09, p= 0.007). Conversely, the probability of full vaccination decreases if the caregiver’s husband exerts control over her decision-making ability (B= -0.29, β = -0.20, p<0.001), or if the caregiver perceives vaccines to be of low importance (B= -0.37, β= -0.27, p<0.001). Belief in religious protection (B= -0.07, β= -0.05, p=0.118) and a belief that vaccines are harmful (B= -0.12, β= -0.04, p= 0.320) did not have an observed effect on vaccination status. This research suggests that interventions may benefit from that including entire families and communities in their design.

## Introduction

Since the establishment of the Expanded Programme on Immunization (EPI) in 1974, vaccinations have contributed to significant reductions in deaths from preventable childhood diseases in low and middle income countries (1). However in recent years vaccination coverage has plateaued or even decreased in some regions, which jeopardises achieving the Immunization Agenda 2030 goal of reducing mortality and morbidity from vaccine-preventable diseases (2,3). In the World Health Organisation (WHO) Africa region, for instance, it was estimated that in 2019, 9.4 million children were under- or unvaccinated, which risks epidemics of infectious disease (4).

Low demand for vaccination among caregivers of young children contributes to stagnating coverage rates across Africa (5). There are various ways to define demand for vaccination, but UNICEF and the World Health Organisation (WHO) describe it as ‘the actions of individuals and communities to seek, support and/ or advocate for vaccines and vaccination services’ (5). Research on this topic in sub-Saharan Africa proposes that demand for vaccination is informed by family and community priorities and power structures; belief in traditional or religious forms of disease prevention; the exchange of information (including rumours and conspiracy theories) in communities; personal experience of vaccination; and interactions with healthcare systems and providers at the point of delivery (6–18).

The research on vaccine demand to date suggests that many inter-dependent and context-specific factors contribute to uptake of vaccination services (19). Despite this, quantitative analyses of determinants of demand or uptake of vaccination are rarely based on an underlying theory, may use single variables as proxies for complex and multidimensional factors, and often use statistical models that do not consider the relationships between constructs that drive demand for vaccination in the real world. For example, as Degarege et al. have pointed out, studies of demand for routine vaccination in India typically assume direct relationships between individual sociodemographic, environmental and psychological variables and the endpoint in logistic regression models (20).

Research which uses an evidence-based theoretical model of vaccine demand and statistical methods that can account for the multi-faceted determinants of demand and complex relationships between them is required to better understand this topic (21). Consequently, the aims of this study were to i) propose a theoretical model for vaccination demand based on published literature and formative qualitative research, ii) use data from quantitative surveys of caregivers of young children to test the overall fit of the model to the theory, and iii) understand the comparative importance of predictors of vaccine demand.

## Methods

### Setting

The research was conducted in Nigeria, Uganda and Guinea, which were chosen to represent African countries with a range of vaccination coverage rates. Among the three, Guinea has the lowest coverage (23.9%) of the basic vaccines recommended by the Expanded Programme on Immunization (EPI), which are the Bacillus Calmette Guerin vaccine for TB, three doses of DTP-HepB-Hib against diphtheria, tetanus, pertussis, Hepatitis B and Haemophilus influenzae b, three doses of oral polio and one dose of measles (22). Nigeria’s coverage was reported at 31.3% and Uganda has the highest coverage among the study geographies, at 55.2% (23,24). In an analysis of Demographic and Health (DHS) vaccination coverage surveys, Guinea had the lowest percentage of fully vaccinated children of the 25 countries included, Nigeria ranked 22/25 and Uganda 16/25 (25).

### Data Collection

Data were collected using a questionnaire (S2 File), designed using the results of a formative qualitative study (19) and a literature review. The questionnaire collected demographic data, household income, and the vaccination status of the participant’s child, as well as perceptual information on their family and community relationships, traditional and religious beliefs, methods of child protection and attitudes to vaccination and vaccination services. The survey contained attitude statements on these topics, to which participants indicated their agreement or disagreement using a 5-point Likert scale. The questionnaire was translated into Yoruba, Hausa and Igbo in Nigeria; Luganda, Runyankole, Samia, Japadhola and Acholi in Uganda; and French in Guinea, so that enumerators could interpret the questions into Malinké, Soussou or Peul, as required. The survey was administered by trained enumerators using Computer Assisted Personal Interviewing (CAPI) devices. Enumerators were trained over the course of four days in each country.

The research was conducted in six states in Nigeria (Lagos, Kano, Enugu, Sokoto, Nasarawa and Rivers), five regions in Uganda (Acholi, Bukedi, Kampala, North Central and Ankole) and five regions in Guinea (Boké, Conakry, N’Zérékoré, Mamou and Kankan). The regions were selected non-randomly with in-country stakeholders (including EPI and government representatives) to include a range of cultural groups and vaccination coverage rates. A multi-stage, stratified sampling methodology was used in each of the regions to select households for interview. Details are given in the Supplementary Materials (S1 File) as the exact procedure varied by country. In general, the sample was stratified by urban or rural setting within each region. Lower-level geographic areas were selected within each stratum and a starting point determined. Households were then selected following a random walk procedure, a household census was taken, and eligible respondents were selected (using a Kish grid if more than one was present). Participants were eligible if they had primary responsibility for the care of a child between 1 and 3 years old. Both male and female participants were eligible for inclusion.

Written informed consent was secured from all participants. An honorarium was provided in the form of a small household item in Nigeria (approximate value of 1000 NGN/ 2.40 USD) and Uganda (5,300 UGX/ 1.50 USD) and in cash in Guinea (369,000 FG/ 40 USD). The study protocol received approval from Makerere University College of Health Sciences Review Board in Uganda (Ref: 724), the National Health Research Ethics Committee of Nigeria (Approval number: NHREC/01/01/2007-25/09/2019) and the Comité Nationale d’Ethique pour la Recherche en Santé in Guinea (Ref: 026/CNERS/20).

### Analysis

Analyses were carried out in R v.4.0.2 using the psych and lavaan packages (26,27). The data and analysis scripts are available in a Github repository [link: https://github.com/jamesbell1991/Vaccines_Structural_Equation_Modelling].

The structural equation modelling process broadly followed the protocol detailed by Schumacker and Lomax (28). Firstly, a factor analysis was conducted on several Likert-scale questions in the survey. The approach was a combination of exploratory analysis (in that no definite factor structure was predetermined) and confirmatory analysis (in that variables were grouped together in themes in advance of the analysis) as described by Kang et al (29). The final factors were determined through an examination of scree plots and factor loadings to produce six factors. Variables with a factor loading less than 0.3 were considered a poor fit.

The composition of these factors and the theoretical basis for including them was arrived at using the results of a literature review and previous qualitative research (Table 1).

**Table 1:**
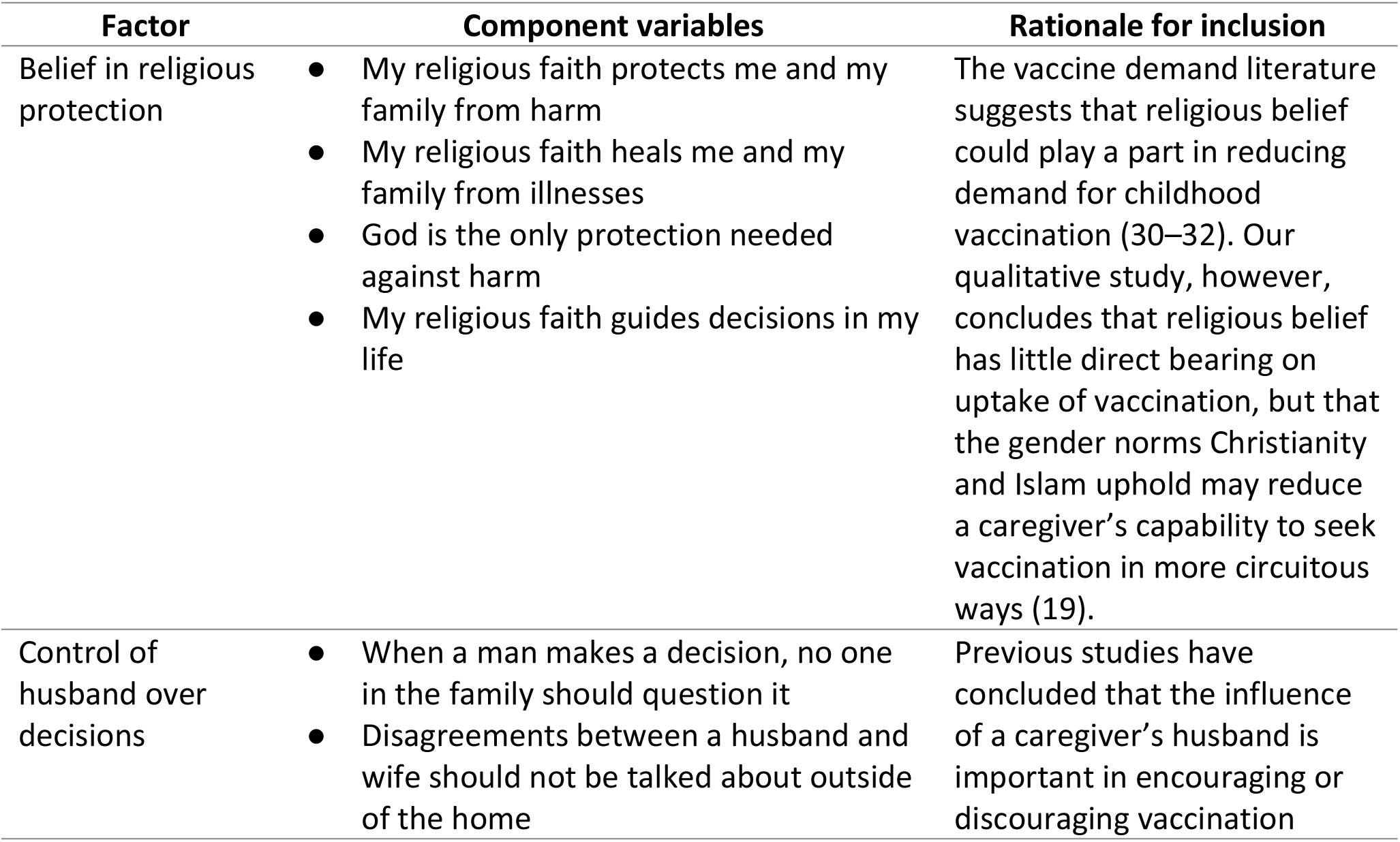
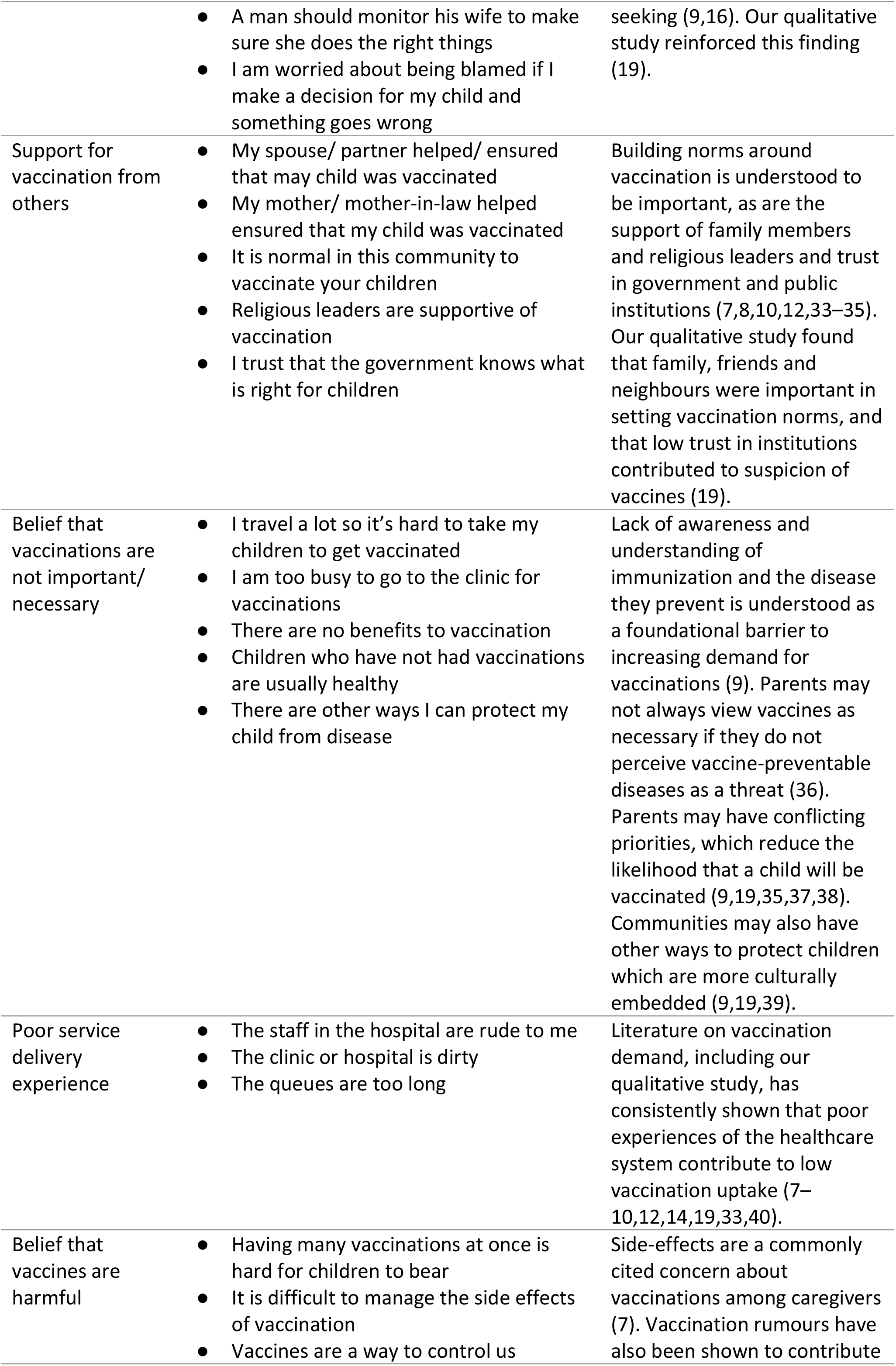
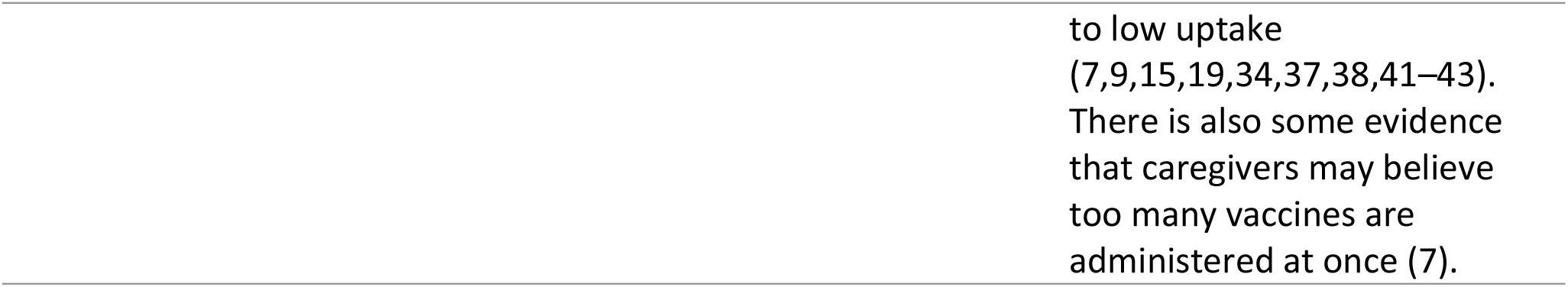
Factor structure and their theoretical justifications

Using these six factors, a structural equation model structure was developed with children’s vaccination status as the dependent/outcome variable. Vaccination status was determined using an adapted version of the protocol used by DHS (44). If available, the vaccines a child had received were determined using the child’s vaccination card. If not available, status was determined by parental reporting, which was not otherwise verified (e.g., through clinic records). For the analysis a dichotomous variable was created to compare children who have completed the full schedule (taking a value of 1) or who have had no doses or some doses but not enough to complete the full schedule (taking a value of 0).

As shown in Figure 1, it was hypothesized that each factor had a direct relationship with the outcome. Existing literature and our previous qualitative study do not support any hypothesized relationships between the factors.

**Fig 1:**
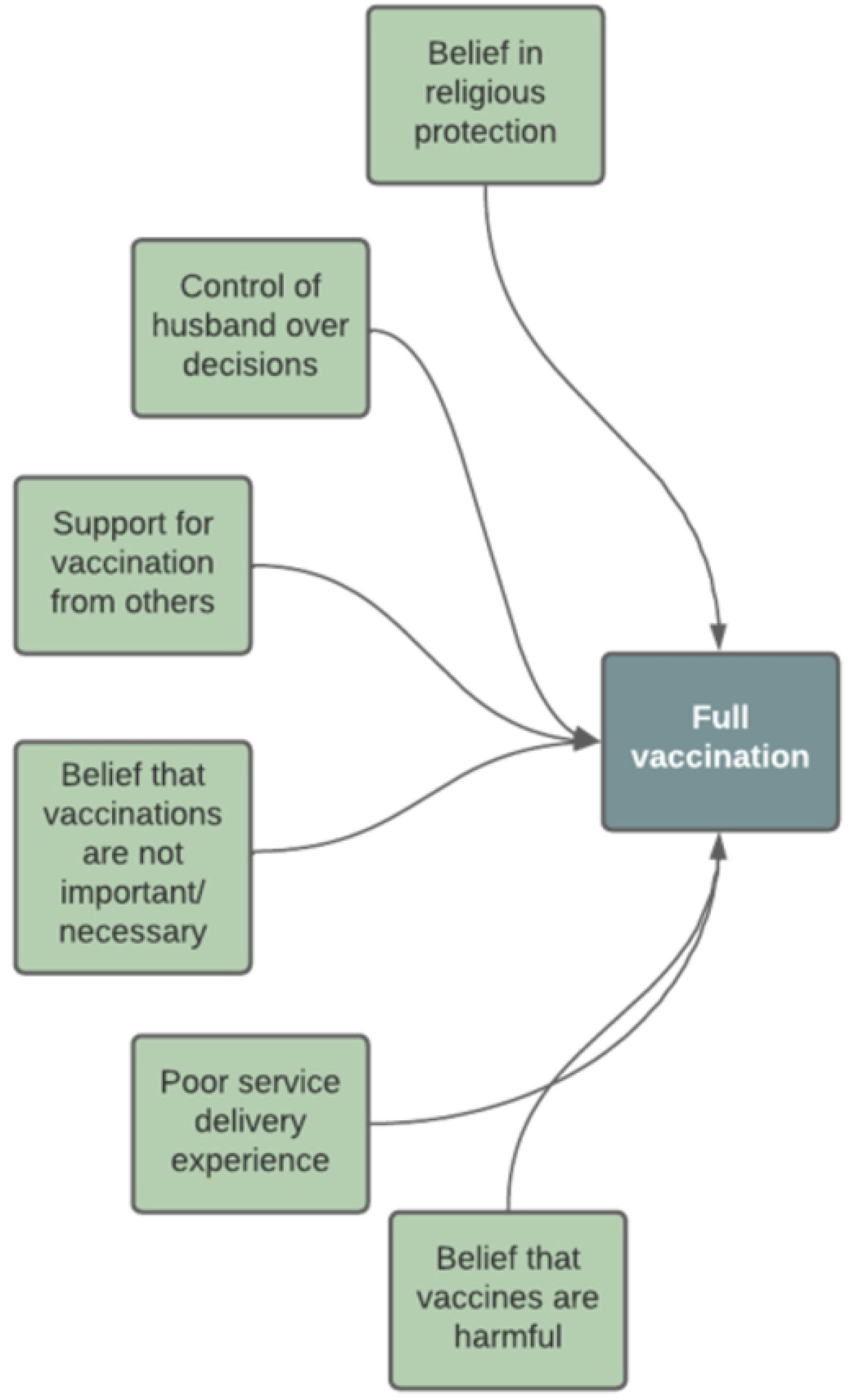
Proposed model structure

The model used a probit link function, to account for the dichotomous outcome variable. Modification indices were examined and additions or deletions to the model were considered. Several goodness of fit indices were examined. No definitive cut-off points were adopted, but the guidance that RMSEA <0.08, TLI >0.90, CFI >0.90 and SRMR <0.08 indicate acceptable fit was used (45). A χ^2^ test was not included due to its sensitivity to sample size (28). Finally, as the countries involved in the study may be heterogenous, models were run for each country separately, the results of which are given in the Supplementary Materials (S3 File).

All tables and figures presented contain sample statistics and have not been weighted to population data.

## Results

### Description of study participants

A total of 3,318 interviews were completed. These took place in Nigeria and Uganda between November and December 2020 and in Guinea between July and August 2021 (later due to resource constraints which prevented the three surveys from running concurrently). Just under a third of interviews were conducted in rural areas (Table 2). Most participants (78*.8%) were under the age of 35. Education levels varied by country: in Nigeria, 56.5% of participants had secondary or higher education, whereas in Uganda most participants had primary education (55.1%). In Guinea, 52.2% of participants had no formal education, the highest of the three countries. In all countries most participants were in the low-income band (see note to Table 2 for definition). 96.0% of participants were the child’s biological mother. Vaccination status of the sample varied by country, with Uganda reporting 60.4% of children fully vaccinated, and lower percentages in Nigeria and Guinea (36.1% and 40.0%, respectively).

**Table 2:** Description of study sample

|  | **Nigeria**<br>**(N=1264)** | **Uganda**<br>**(N=1054)** | **Guinea**<br>**(N=1000)** | **Total**<br>**(N=3318)** |
| --- | --- | --- | --- | --- |
| **Setting** |  |  |  |  |
| Urban | 489 (38.7%) | 406 (38.5%) | 363 (36.3%) | 1258 (37.9%) |
| Rural | 775 (61.3%) | 648 (61.5%) | 637 (63.7%) | 2060 (62.1%) |
| **Age** |  |  |  |  |
| 18-24 | 261 (20.6%) | 358 (34.0%) | 262 (26.2%) | 881 (26.6%) |
| 25-29 | 406 (32.1%) | 312 (29.6%) | 340 (34.0%) | 1058 (31.9%) |
| 30-34 | 268 (21.2%) | 191 (18.1%) | 214 (21.4%) | 673 (20.3%) |
| 35-39 | 216 (17.1%) | 124 (11.8%) | 131 (13.1%) | 471 (14.2%) |
| 40-44 | 83 (6.6%) | 48 (4.6%) | 33 (3.3%) | 164 (4.9%) |
| 45-49 | 26 (2.1%) | 15 (1.4%) | 16 (1.6%) | 57 (1.7%) |
| 50-56 | 4 (0.3%) | 6 (0.6%) | 4 (0.4%) | 14 (0.4%) |
| **Education** |  |  |  |  |
| No formal education | 387 (30.6%) | 74 (7.0%) | 522 (52.2%) | 983 (29.6%) |
| Primary | 163 (12.9%) | 581 (55.1%) | 185 (18.5%) | 929 (28.0%) |
| Secondary | 530 (41.9%) | 319 (30.3%) | 188 (18.8%) | 1037 (31.3%) |
| Higher education | 184 (14.6%) | 79 (7.5%) | 77 (7.7%) | 340 (10.2%) |
| Prefer not to answer | 0 (0%) | 1 (0.1%) | 28 (2.8%) | 29 (0.9%) |
| **Income level** |  |  |  |  |
| Low | 730 (57.8%) | 759 (72.0%) | 543 (54.3%) | 2032 (61.2%) |
| Middle | 306 (24.2%) | 228 (21.6%) | 115 (11.5%) | 649 (19.6%) |
| High | 131 (10.4%) | 24 (2.3%) | 2 (0.2%) | 157 (4.7%) |
| Prefer not to say | 97 (7.7%) | 43 (4.1%) | 340 (34.0%) | 480 (14.5%) |
| **Number of children** |  |  |  |  |
| Mean (SD) | 2.92 (1.89) | 3.35 (2.29) | 3.69 (2.18) | 3.29 (2.13) |
| Median [Min, Max] | 3.00 [1.00, 20.0] | 3.00 [1.00, 20.0] | 3.00 [1.00, 16.0] | 3.00 [1.00, 20.0] |
| **Relationship to child** |  |  |  |  |
| Biological mother | 1195 (94.5%) | 1038 (98.5%) | 953 (95.3%) | 3186 (96.0%) |
| Stepmother | 18 (1.4%) | 4 (0.4%) | 3 (0.3%) | 25 (0.8%) |
| Aunt | 40 (3.2%) | 7 (0.7%) | 16 (1.6%) | 63 (1.9%) |
| Grandmother | 3 (0.2%) | 4 (0.4%) | 0 (0%) | 7 (0.2%) |
| Biological father | 7 (0.6%) | 1 (0.1%) | 4 (0.4%) | 12 (0.4%) |
| Stepfather | 1 (0.1%) | 0 (0%) | 2 (0.2%) | 3 (0.1%) |
| Other | 0 (0%) | 0 (0%) | 22 (2.2%) | 22 (0.7%) |
| **Child's vaccination status** |  |  |  |  |
| Not vaccinated | 143 (11.3%) | 87 (8.3%) | 106 (10.6%) | 336 (10.1%) |
| Partially vaccinated | 665 (52.6%) | 330 (31.3%) | 494 (49.4%) | 1489 (44.9%) |
| Fully vaccinated | 456 (36.1%) | 637 (60.4%) | 400 (40.0%) | 1493 (45.0%) |

Note: Income bands per country (per month). Low: Nigeria (Below 50,000 NGN), Uganda (Below 500,000 UGX), Guinea (Below 1,983,626 GNF); Middle: Nigeria (50,0001-500,000 NGN), Uganda (501,000-2,000,000 UGX), Guinea (1,983,627-4,999,999 GNF); High: Nigeria (Above 800,0001 NGN), Uganda (Above 2,000,000 UGX), Guinea (Above 5,000,000 GNF)

### Measurement Model

The measurement model corresponding to the six latent factors in Table 1 fitted the data reasonably well (RMSEA = 0.04, TLI = 0.88, CFI = 0.89, SRMR = 0.04). To improve the fit further, we allowed some residual terms within the same construct to covary (My spouse/ partner helped/ ensured that my child was vaccinated with My mother/ mother-in-law helped/ ensured that my child was vaccinated and It is normal in this community to vaccinate your children with Religious leaders are supportive of vaccination), resulting in the final measurement model (Table 3). One variable (Disagreements between a husband and wife are private and should not be talked about outside the home) had a standardised factor loading of 0.294 but was retained in the model as its removal did not appreciably improve the fit statistics.

**Table 3:** Measurement Model Factor Loadings and Fit Statistics

| Factor | Variable | Factor loading | Standard error | P-value | Standardised factor loading |
| --- | --- | --- | --- | --- | --- |
| **Belief in religious protection** | My religious faith protects me and my family from harm | 1.000 |  |  | 0.755 |
|  | My religious faith heals me and my family from illnesses | 1.196 | 0.037 | <0.001 | 0.708 |
|  | God is the only protection needed against harm | 0.617 | 0.023 | <0.001 | 0.553 |
|  | My religious faith guides decisions in my life | 0.710 | 0.024 | <0.001 | 0.612 |
| **Control of husband over decisions** | When a man makes a decision, no one in the family should question it | 1.000 |  |  | 0.498 |
|  | A man should monitor his wife to make sure she does the right things | 0.827 | 0.059 | <0.001 | 0.522 |
|  | Disagreements between a husband and wife are private and should not be talked about outside the home | 0.474 | 0.045 | <0.001 | 0.294 |
|  | I am worried about being blamed if I make a decision for my baby/ child and something goes wrong | 0.542 | 0.050 | <0.001 | 0.301 |
| **Support for vaccination from others** | My spouse / partner helped/ ensured that my child was vaccinated | 1.000 |  |  | 0.542 |
|  | My mother/ mother-in-law helped/ ensured that my child was vaccinated | 1.045 | 0.045 | <0.001 | 0.512 |
|  | It is normal in this community to vaccinate your children | 0.778 | 0.037 | <0.001 | 0.612 |
|  | Religious leaders are supportive of vaccination | 0.747 | 0.040 | <0.001 | 0.485 |
|  | I trust that the government knows what is right for children | 0.891 | 0.045 | <0.001 | 0.541 |
| **Belief that vaccinations are not important/ necessary** | I travel a lot so it's hard to take my child to get vaccinated | 1.000 |  |  | 0.528 |
|  | I am too busy to go to the clinic or hospital for vaccinations | 1.115 | 0.047 | <0.001 | 0.528 |
|  | There are no benefits to vaccination | 1.123 | 0.058 | <0.001 | 0.561 |
|  | Children who have not had vaccinations are usually healthy | 1.059 | 0.058 | <0.001 | 0.497 |
|  | There are other ways I can protect my child from disease | 0.732 | 0.052 | <0.001 | 0.334 |
| **Poor service delivery experience** | The staff in the hospital are rude to me | 1.000 |  |  | 0.612 |
|  | The clinic or hospital is dirty | 1.029 | 0.058 | <0.001 | 0.664 |
|  | The queues are too long at the clinic/ hospital where the vaccination takes place | 0.455 | 0.035 | <0.001 | 0.309 |
| **Belief that vaccines are harmful** | Having many vaccinations at once is hard for children to bear | 1.00 |  |  | 0.356 |
|  | It is difficult for me to manage the side effects (fever, rash, pain) of vaccination | 1.843 | 0.139 | <0.001 | 0.586 |
|  | Vaccines are a way for global/western countries/organisations to control us | 1.682 | 0.128 | <0.001 | 0.530 |
| RMSEA = 0.04, TLI = 0.90, CFI = 0.91, SRMR = 0.04 |  |  |  |  |  |

### Structural Model

The fit statistics for the model indicate acceptable model fit: RMSEA = 0.04, TLI = 0.91, CFI = 0.92, SRMR = 0.04. Modification indices were examined, but none were logical within the theoretical framework so none were adopted.

### Factors affecting uptake of childhood vaccination

Some factors are associated with a reduction in the probability that a child would be vaccinated, while others lead to an observed increase in the probability of vaccination, and others were unassociated with the outcome. (Table 4).

**Table 4:** Unstandardised (B) and standardised (β) effects of factors affecting vaccination in the structural model

| Factor | B (95% CI) | $\beta$ (95% CI) | P-value |
| --- | --- | --- | --- |
| Belief in religious protection | -0.07 (-0.17, 0.02) | -0.05 (-0.11, 0.01) | 0.118 |
| Control of husband over decisions | -0.29 (-0.43, -0.14) | -0.20 (-0.29, -0.11) | <0.001 |
| Support for vaccination from others | 0.33 (0.19, 0.46) | 0.21 (0.13, 0.30) | <0.001 |
| Belief that vaccinations are not important/ necessary | -0.37 (-0.51, -0.22) | -0.27 (-0.37, -0.17) | <0.001 |
| Poor service delivery experience | 0.09 (0.02, 0.16) | 0.09 (0.03, 0.16) | 0.007 |
| Belief that vaccines are harmful | -0.12 (-0.37, 0.12) | -0.04 (-0.13, 0.04) | 0.320 |

Lower probability of vaccination was observed for those who expressed higher levels of perceived control of the husband over decision-making (B -unstandardised effect= -0.29, β-standardised effect = -0.20, p<0.001). The unstandardised effect can be interpreted to mean that when this variable increases by one unit, the z-score for probability of being fully vaccinated decreases by 0.29 units. The standardised coefficient can be interpreted to mean that when this variable is increased by one standard deviation, the z-score score for probability of being fully vaccinated decreases by 0.20 standard deviations. Lower probability was also observed for those who expressed higher levels of belief that vaccinations are not important or necessary (B= -0.37, β= 0.27, p<0.001). Higher probabilities of vaccination were observed for participants who said that they had higher levels of support for vaccination from others around them (B= 0.33, β= 0.21, p<0.001) and among those who had worse service delivery experiences (B= 0.09, β= 0.09, p= 0.007). There was little evidence that belief in religious protection (B= -0.07, β= -0.05, p=0.118) or belief that vaccines are harmful (B= -0.12, β= -0.04, p= 0.320) increased or decreased the probability of vaccination.

In a comparison of the standardised coefficients (β), the factor with the strongest positive observed impact on vaccination was having support from others to vaccinate. The strongest negative impacts were observed for those who expressed high degrees of control of decisions by the husband, and stronger beliefs that vaccinations were not important or necessary.

## Discussion

This study used structural equation modelling to examine factors associated with uptake of childhood vaccination among primary caregivers in Uganda, Guinea and Nigeria. The results suggest that vaccination uptake is informed by family and community relationships, service delivery experience and attitudes and beliefs towards vaccination. Elements of the findings were consistent with existing research on this topic. Higher levels of spousal control over decision-making were again linked to lower likelihood to vaccinate, the role of community norms in encouraging vaccination was reaffirmed, and the importance of belief in the necessity of vaccines in the context of other priorities was observed (9,10,12). The study also provides new contributions to our understanding of the determinants of vaccine demand in several ways. Thematically, the study gives alternative perspectives on the role of religious belief and healthcare service experience compared to what is prevalent in the literature. Conceptually, the work departs from standard methodologies employed in vaccine demand research by using analytical approaches that account for the complexity of the factors that inform vaccine uptake, and which are based on underlying data-driven theories of behaviour.

Given what is reported elsewhere in the literature, two of the study’s conclusions may appear surprising. Others have suggested that caregiver belief in religious protection may decrease likelihood of vaccine uptake (30–32). Our findings do not support this hypothesis, which is in line with the results of our qualitative research on the same topic (19). It is possible that religious protection and protection conferred by vaccines are seen as conceptually separate, and with different functions in child development. This means that interventions to increase demand for vaccination should be careful not to attempt to supplant belief in religious protection with a preference for vaccination. Interventions involving religious community leaders (such as have been attempted in Nigeria) could be fruitful avenues to ensure that different conceptions of child protection are viewed as complementary rather than adversarial (46–48).

It is well established that poor service delivery experiences may discourage caregivers from seeking vaccination (7–9,12–14,19). Even though the effect size observed in our study was small, it is surprising that our results suggest that caregivers who experience worse service delivery experience are more likely to have fully vaccinated children. There are several possible explanations for this finding. In the country-level analysis (presented in the Supplementary Materials, S3 File) the association is driven by the data from Guinea, which suggests that the finding may be due to sampling or cognitive biases in questionnaire responses that are specific to that country. Informal conversations with the fieldwork teams revealed that participants were at times unwilling to give negative opinions about the government, which may have affected responses to the variables comprising this factor. Alternatively, it is theoretically plausible that those who had fully vaccinated children are more dissatisfied with the experience of vaccinating at the clinic, compared to those with un- or under-vaccinated children, who will have had fewer touchpoints with health services. Finally, the result could have been the result of uncontrolled confounding by variables that were not included in the model.

Our study’s results also support the idea that vaccination uptake is not determined solely by the attitudes and behaviours of the child’s primary caregiver, but by a range of intersecting familial, community and social influences. This suggests that ‘whole family’ or ‘whole community’ intervention approaches could be impactful in these contexts. Programmes based on principles of collectivism encourage families and communities to adopt a desired behaviour together, and have shown promise in other policy areas and geographies (49,50).

When the analysis is done separately by country, some differences by geography are noted. In Nigeria, support from others is observed to drive vaccination uptake, and bad service delivery impedes it. In Uganda, practical difficulties are the sole barrier to uptake, and in Guinea support from others, bad service delivery and belief in religious protection increase the probability of vaccination and belief that vaccinations are harmful decreases it. These differences mean that interventions should ensure that local contexts are taken into account when designing strategies to encourage adoption of vaccination.

This study moved beyond the standard approach in many explorations of predictors of childhood vaccination demand, which may rely on observed variables only as model inputs. Determinants of demand are often multifaceted in nature, necessitating the use of latent variables or constructs (21). In this way, our study was able to engage with the complexity of the phenomenon more holistically in its analytical approach. In addition, our analysis was also explicitly based on themes identified through prior qualitative research. A research-based approach, and the choice of structural equation modelling as the analytical tool, ensured that the hypothesised relationships between the explanatory factors had an empirical basis and were stated explicitly rather than assumed. This may result in models that reflect more closely how decisions around vaccination play out in the real world, which may make resulting interventions more appropriate.

Further research on this topic could undertake more complex analysis than has been attempted here. This could include developing factors to describe other important constructs that may affect vaccination (such socio-economic status or belief in gender norms), proposing and testing more elaborate relational structures between factors, or the exploration of potential moderation or mediation between latent constructs.

### Limitations

Some important limitations should be considered when evaluating the research findings. All answers were self-reported and not verified using external sources, so the vaccination outcome data may have been over- or under-stated. Attitudinal questions may have been affected by social desirability or recall biases. The sampling methodology should have resulted in regionally representative samples, but the random-walk methodology could have introduced sampling bias (51). The differences between the sampling protocols (as explained the Supplementary Materials) could also reduce comparability between countries.

The factors included in the model were partially determined by the availability of data, and therefore important constructs are likely absent from the analysis, rendering it an incomplete view of the determinants of vaccination uptake.

Finally, the standardised factor loading scores are considered low by many measures, meaning that the cohesiveness of the latent constructs and the regressions based on them are open to critique (52). The decision to combine data from three heterogeneous countries is also open to criticism as it may obscure country-level dynamics (but this is remedied by the inclusion of country-level models in the Supplementary Materials. S3 File).

## Conclusion

Research on vaccination uptake often relies on proxy variables to represent complex phenomena and may not be based on an underlying theory of how vaccination decisions are made. This article uses the results of a formative qualitative study to construct and test a model to help explain determinants of vaccination uptake. We conclude that uptake is informed by family and community relationships, service delivery experience and attitudes and beliefs towards vaccination. The work has implications for intervention design and suggests that approaches that include entire families and communities in interventions may be beneficial.

## Data Availability

All data are available in this repository: https://github.com/jamesbell1991/Vaccines_Structural_Equation_Modelling

## Acknowledgments

We would like to acknowledge the contributions of Virginia Nkwanzi, Alhassane Baldé, Mohamed Dioubaté, Idalecio Agostinho das Neves, Possy Mugyenyi, Lisa Oot, Kate Bagshaw, Rebecca Fields, Ugochukwu Osigwe, Ndadilnasiya Endie Waziri, Yusuf Yusufari, Jenny Sequeira, Sarah Chesemore, Wenfeng Gong, Anna Rapp, Tracy Johnson, Andrew Buhayar and Olesia Savateeva during the fieldwork and analysis phases of the study, and the enumerators and participants involved in the research.

## Supporting information captions

S1 File. Sampling Protocols

S2 File. Questionnaires

S3 File. Country Analysis

## Notes

### Competing Interest Statement

All authors work for market and social research organisations that carry out research on behalf of commercial companies, including in the field of vaccines.

### Funding Statement

This work was supported by the Bill and Melinda Gates Foundation (www.gatesfoundation.org) through a grant received by Ipsos Healthcare, Investment ID INV-001411. All authors received salaries from either Ipsos Healthcare, Ipsos Nigeria, Ipsos Uganda, or Ciblage to complete the work. The funder provided input into study design, data analysis, and the final manuscript, but all final decisions were taken by members of the study team.

### Author Declarations

The study protocol received approval from Makerere University College of Health Sciences Review Board in Uganda (Ref: 724), the National Health Research Ethics Committee of Nigeria (Approval number: NHREC/01/01/2007-25/09/2019) and the Comité Nationale d'Ethique pour la Recherche en Santé in Guinea (Ref: 026/CNERS/20).

